# Machine Learning for Sudden Cardiac Death Prediction in the Atherosclerosis Risk in Communities Study

**DOI:** 10.1101/2022.01.12.22269174

**Authors:** Zhi Yu, Shannon Wongvibulsin, Natalie R. Daya, Linda Zhou, Kunihiro Matsushita, Pradeep Natarajan, Josef Coresh, Scott L. Zeger

## Abstract

**Introduction:** Sudden cardiac death (SCD) is a devastating consequence often without antecedent expectation. Current risk stratification methods derived from baseline independently modeled risk factors are insufficient. Novel random forest machine learning (ML) approach incorporating time-dependent variables and complex interactions may improve SCD risk prediction.

**Methods:** Atherosclerosis Risk in Communities (ARIC) study participants were followed for adjudicated SCD. ML models were compared to standard Poisson regression models for interval data, an approximation to Cox regression, with stepwise variable selection. Eighty-two time-varying variables (demographics, lifestyle factors, clinical characteristics, biomarkers, etc.) collected at four visits over 12 years (1987-98) were used as candidate predictors. Predictive accuracy was assessed by area under the receiver operating characteristic curve (AUC) through out-of-bag prediction for ML models and 5-fold cross validation for the Poisson regression models.

**Results:** Over a median follow-up time of 23.5 years, 583 SCD events occurred among 15,661 ARIC participants (mean age 54 years and 55% women). Compared to different Poisson regression models (AUC at 6-year ranges from 0.77-0.83), the ML model improved prediction (AUC at 6-year 0.89). Top predictors identified by ML model included prior coronary heart disease, which explained 47.9% of the total phenotypic variance, diabetes mellitus, hypertension, and T wave abnormality in any of leads I, aVL, or V6. Using the top ML predictors to select variables, the Poisson regression model AUC at 6-year was 0.77 suggesting that the non-linear dependencies and interactions captured by ML, are the main reasons for its improved prediction performance.

**Conclusions:** Applying novel ML approach with time-varying predictors improves the prediction of SCD. Interactions of dynamic clinical characteristics are important for risk-stratifying SCD in the general population.

## Introduction

Sudden cardiac death (SCD) accounts for approximately 400,000 adult deaths in the United States each year.^1-3^ Risk stratification for SCD continues to lag contributing to the significant public health burden.^4^ Current guidelines directing the use of primary prevention, implanted cardioverter defibrillator (ICD), largely rely on a single parameter: reduced left ventricular ejection fraction (LVEF). Using LVEF alone is inadequate for two main reasons: (1) it does not account for dynamic factors including interim clinical events;^5^ and (2) it is seldom measured for approximately 50% of SCD victims who did not have a prior diagnosis of heart disease.^1,4^ Previous efforts to expand the list of SCD predictors examined single or small numbers of static predictors, or were conducted among patients with known existing cardiovascular disease.^6-11^

By combining data from a general population cohort with dense phenotyping and a novel machine learning (ML) approach capable of handling large number of time-dependent variables and incorporating non-linear relationships as well as complex interactions between risk factors, we aimed to develop a population-based approach to identify individuals at high SCD risk.

## Methods

### Study Cohort

The Atherosclerosis Risk in Communities (ARIC) study is an ongoing longitudinal cohort of 15,792 middle-aged men and women recruited from four communities in the U.S.: Forsyth County, North Carolina; Jackson, Mississippi; suburbs of Minneapolis, Minnesota; and Washington County, Maryland at 1987-1989 (visit 1). The 3 short-term follow-up visits occurred approximately three years apart: 1990-1992 (visit 2), 1993-1995 (visit 3), 1996-1998 (visit 4). Each study visit consisted of a comprehensive examination, which included physical and clinical examination, blood and urine specimens’ collection for laboratory testing, administration of extensive questionnaires, and a 12-lead electrocardiogram (ECG). In addition, participants were contacted by phone annually for hospitalizations and death information during the prior year. If any clinical events happened, hospital records and death certificates would be obtained for ascertainment by physicians.^12^ In the current study, participants with ICD (N=131) were excluded from the analysis. Study protocols were approved by the Institutional Review Boards and all study participants provided informed consent.

### Assessment of Candidate Predictors

Clinical predictors were measured and updated during the first four ARIC visits from 1987-89 to 1996-98. At visit 1, demographics variables including age at the time of the visit, sex, race, center, education level, income, and family history (mother and father) of diseases were collected using an interviewer-administered questionnaire. Anthropometric variables including height, weight, and waist circumstances were measured by standard protocol at all visits, and body mass index (BMI) was calculated as weight (in kilograms) divided by the square of height (in meters). Lifestyle factors including smoking status, alcohol intake, physical activity [intensity and meeting American Heart Association (AHA) recommendations or not], and dietary quality were evaluated using questionnaires that assess each participant’s self-reported information at all visits (alcohol intake and smoking status) or visit 1 and 3 (physical activity and dietary quality).^13-16^ Clinical factors included systolic and diastolic blood pressure, hypertension, diabetes mellitus, coronary heart disease (CHD), stroke, atrial fibrillation, heart failure, and hospitalizations (yes/no and number of hospitalizations). These variables were evaluated at all visits.

The definitions and ascertainment of the clinical factors are described as follows. Blood pressure was measured in seated participants after a 5-minute rest. Hypertension was defined as systolic blood pressure ≥ 140 mm Hg, diastolic blood pressure ≥ 90 mm Hg, or use of antihypertensive medication in the 2 weeks prior to visits.^17^ Diabetes mellitus was defined as fasting blood glucose ≥ 126 mg/dL, non-fasting glucose ≥ 200 mg/dL, self-reported doctor-diagnosed diabetes, or use of diabetes medication in the 2 weeks prior to visits. CHD, at visit 1, was defined as myocardial infarction (MI) observed on ECG, self-reported history of MI, self-reported heart or arterial surgery, coronary bypass, balloon angioplasty, and coronary angioplasty and, at visit 2-4, additionally included CHD cases occurring after visit 1 but before the relevant visit which were identified based on hospitalization records and death records and adjudicated by physicians. Stroke, at visit 1, was defined self-reported stroke and, at visit 2-4, additionally included stroke cases occurring after visit 1 but before the relevant visit which were identified by a computer diagnostic algorithm detailed elsewhere and adjudicated by physicians.^18^ Atrial fibrillation was identified through ECGs from follow-up exams, hospital discharge records, and death certificates. Heart failure, at visit 1, was defined as use of heart failure medication or evidence of symptoms defined by stage 3 of the Gothenburg criteria and, at visit 2-4, additionally included heart failure cases occurring between visit 1 and the relevant visit which were identified though ICD codes in hospitalization and death records.^19^ For clinical events including diabetes, CHD, stroke, atrial fibrillation, and heart failure, incidences were only considered until visit 4. All-cause hospitalizations, cardiac-related hospitalization, and pulmonary-related hospitalization were ascertained through hospitalization records obtained from annual telephone contact with participants and active surveillance in the study community hospitals. Use of medication, such as anti-hypertensive, anti-arrhythmic, lipid-lowering, anti-diabetic medication, were obtained by extracting medication names or codes from the lists transcribed from the medication containers study participants brought to the study visits.

Laboratory values or biomarkers included C-reactive protein (CRP), white blood cells (WBC) count, hematocrit, hemoglobin, N-terminal pro-brain natriuretic peptide (NT-proBNP), troponin I, troponin T, fibrinogen, total cholesterol, high-density lipoprotein cholesterol (HDL-C), low-density lipoprotein cholesterol (LDL-C), triglycerides, serum creatinine, estimated glomerular filtration rate (eGFR), serum albumin, urine albumin, blood glucose (fasting and non-fasting), serum sodium, serum potassium, blood urea nitrogen, and serum magnesium. Definition of each marker and at which visit(s) each marker was measured are described in detail in the **Supplemental Table 1**.

**Table 1.** Baseline characteristics of Atherosclerosis Risk in Communities (ARIC) study participants.

| Metric* | No SCD (N = 15,078) | SCD (N = 583) | P-value |
| --- | --- | --- | --- |
| **Demographics & anthropometric variables** |  |  |  |
| Age (years) | 54.1 (5.8) | 56.2 (5.6) | <0.001 |
| Male | 6623 (43.9) | 365 (62.6) | <0.001 |
| White | 11048 (73.5) | 338 (58.0) | <0.001 |
| At least high school degree | 11546 (76.7%) | 358 (61.5%) | <0.001 |
| Body mass index (kg/m<sup>2</sup>) | 27.7 (5.3) | 29.3 (6.0) | <0.001 |
| **Lifestyle factors** |  |  |  |
| Current smoker | 3886 (25.8) | 216 (37.0) | <0.001 |
| Current alcohol drinker | 8422 (56.1) | 273 (47.2) | <0.001 |
| Physical activity (MET-min/week) | 611.8 (764.7) | 502.2 (684.1) | 0.001 |
| **Clinical factors** |  |  |  |
| Systolic blood pressure (mmHg) | 120.9 (18.7) | 131.0 (22.6) | <0.001 |
| Diastolic blood pressure (mmHg) | 73.6 (11.2) | 76.9 (13.3) | <0.001 |
| Hypertension | 5086 (33.9) | 351 (60.6) | <0.001 |
| Coronary heart disease | 600 (4.1) | 133 (23.5) | <0.001 |
| Heart failure | 683 (4.6) | 61 (10.7) | <0.001 |
| Stroke | 239 (1.6) | 43 (7.4) | <0.001 |
| Diabetes mellitus | 1089 (7.2) | 131 (22.5) | <0.001 |
| **Medication classes** |  |  |  |
| Anti-hypertensive | 4455 (29.6) | 319 (54.7) | <0.001 |
| Lipid-lowering | 423 (2.8) | 23 (4.0) | 0.129 |
| Anti-diabetic | 774 (6.1) | 100 (19.6) | <0.001 |
| Anti-arrhythmic | 104 (0.7) | 14 (2.4) | <0.001 |

| **Laboratory values or biomarkers** |  |  |  |
| --- | --- | --- | --- |
| HDL-C, md/dL | 49.1 [39.5, 61.6] | 43.3 [35.6, 53.7] | <0.001 |
| LDL-C, md/dL | 134.9 [110.8, 160.8] | 146.3 [119.3, 174.9] | <0.001 |
| Triglyceride, md/dL | 109.0 [78.0, 156.0] | 124.0 [88.0, 188.8] | <0.001 |
| Total cholesterol, md/dL | 212.0 [186.0, 239.0] | 219.0 [193.0, 251.0] | <0.001 |
| eGFR, mL/min/1.73 m<sup>2</sup> | 102.6 (15.7) | 99.6 (19.2) | <0.001 |
| **Electrocardiogram variables** |  |  |  |
| Atrial fibrillation | 34 (0.2) | 3 (0.5) | 0.324 |
| Heart rate, beats per minute | 66.7 (10.3) | 67.8 (11.5) | 0.015 |
| Presence of diagnostic Q wave | 131 (0.9) | 44 (7.7) | <0.001 |
| Cornell voltage (S in V3+R in aVL), uV | 1245.8 (556.6) | 1556.1 (650.1) | <0.001 |
| Left ventricular hypertrophy | 308 (2.1) | 33 (5.9) | <0.001 |

\*Values are expressed as mean (SD) or median [IQR] for continuous variables and N (%) for categorical variables.
eGFR: Estimated glomerular filtration rate; HDL-C: high-density lipoprotein cholesterol; LDL-C: low-density lipoprotein cholesterol.

ECG variables were derived based on the 12-lead ECGs results measured at all visits. Details of the ECG measuring protocol, data processing, monitoring, and quality control have been described elsewhere.^20^ Twenty-one variables of ECG patterns classified by Minnesota Code, which utilized a series of measurement rules to assign numerical codes based on the location and severity of ECG findings, were included.^21,22^ The presence of diagnostic Q-wave, QRS duration, QT duration, and heart rate observed from ECG were also used as candidate predictors. Cornell voltage (S in V3+R in aVL) calculated from ECG results for evaluating left ventricular thickness, as well as left ventricular hypertrophy (LVH) defined by Cornell voltage criteria.^23^

### Assessment of Outcome

SCD was defined as a sudden pulseless condition presumed due to a ventricular tachyarrhythmia in a previously stable individual without evidence of a non-cardiac cause of cardiac arrest occurring out of the hospital or in the emergency department. Details regarding the ascertainment of SCD events in ARIC have been described elsewhere.^24^ Briefly, all fatal CHD events documented in death certificates, informant interviews, physician questionnaires, coroner reports, prior medical history, or hospital discharge summaries that occurred through 2012 were reviewed and independently adjudicated by 2 physicians. Cases were classified as definite sudden arrhythmic death, possible sudden arrhythmic death, not sudden arrhythmic death, or unclassifiable. For the current analysis, we defined SCD as adjudicated definite or possible sudden arrhythmic death that occurred by December 31, 2012.

### Statistical Analysis

We used the Random Forest for Survival, Longitudinal, and Multivariate data analysis (RF-SLAM) methods for SCD risk prediction. Details of this method have been described elsewhere.^25^ Briefly, random forest is an ensemble learning method that combines multiple decision trees trained on uncorrelated bootstrap replications of the original training data and outputs the averaged prediction across all trees.^26^ RF-SLAM uses data partitioned into discrete intervals (also known as person-time intervals) and uses a Poisson regression log-likelihood as the split statistic thus allowing for modeling time-varying predictor variables. In this analysis, we followed the recommended settings of 1,000 as the number of trees, 10% of the total number of intervals as the minimum terminal node size, and the square root of the number of input variables as the number of predictors for each tree.^25^ To simplify the interpretation of the RF-SLAM predictions, we grew a single regression tree that best approximates the RF-SLAM predictions among those that explained at least 90% of the variation in the RF-SLAM predictions.^27^ We grew a second tree that explained at least 80% of the variation for facilitating visualization and discussion of key predictors and their interactions.

We compared the performance of RF-SLAM to a series of standard Poisson regression models that share the interval event indicator as the outcome and differ by their specific variable selection approach. Missing data was handled through imputation by randomly drawing from non-missing data by RF-SLAM and by multivariate imputation by chained equations for the Poisson regression models.^28-30^ The RF-SLAM and the Poisson regression models shared the same pool of 82 potential (dynamic) predictor variables collected at the four visits including demographics, anthropometric variables, lifestyle factors, clinical characteristics, medication, laboratory values and biomarkers, ECG variables, and other cardiac functional indices. To evaluate the importance of time-varying covariates, we additionally include a random forest survival (RFS) model incorporating only baseline covariates.^31^

We considered the following pools of predictor variables for the Poisson regressions: (1) using the top predictors as selected in the summary regression tree that explained at least 80% of the variation of RF-SLAM (referred to as top predictors of RF-SLAM); (2) using top predictors of RF-SLAM as candidate predictors with selection by stepwise regression, (3) using top predictors of RF-SLAM plus interactions formed by parent-child node pairs identified from the summary regression tree of RF-SLAM and modeled based on the tree structure (referred to as top interactions of RF-SLAM) as predictors, and (4) using top predictors of RF-SLAM plus top interactions of RF-SLAM as candidate predictors with selection by stepwise regression. Predictive accuracy was assessed at 3, 6, 9, and 25 years by time-dependent area under the receiver operating characteristic curve (AUC) through out-of-bag prediction for RF-SLAM and RFS models and 5-fold cross validation for Poisson regression models.^32^ The comparison between the RF-SLAM model, RFS model, and Poisson regression models is also summarized in **Supplemental Table 2**. Since multiple variables were measured in only one or two out of the total four visits resulting in substantial missingness we excluded all variables with greater than 50% missingness in sensitivity analysis to test the robustness of our results. Also, as CHD appeared to be the key predictor of SCD in our main model, as shown below, we excluded all participants with existing CHD at baseline as a sensitivity analysis. Analyses were conducted with R version 3.6.3 (https://www.r-project.org). Statistical significance was defined as *P* value < 0.05.

**Table 2.** Predictors that together explained > 90% of the RF-SLAM prediction for sudden cardiac death.

|  | **Variable** | **Category** | **% of total variation explained** |
| --- | --- | --- | --- |
| 1 | Prior coronary heart disease* | Clinical factors | 47.95% |
| 2 | Diabetes mellitus* | Clinical factors | 12.29% |
| 3 | Hypertension* | Clinical factors | 4.97% |
| 4 | N-terminal pro-brain natriuretic peptide* | Laboratory values or biomarkers | 3.78% |
| 5 | T wave abnormality in any of leads I, aVL, and V6* | Electrophysiologic variables | 3.61% |
| 6 | Number of cardiac-related hospitalization* | Clinical factors | 2.39% |
| 7 | Use of anti-diabetic medications* | Medications | 2.13% |
| 8 | ST junction & segment depression in any of leads I, aVL, or V6* | Electrophysiologic variables | 1.47% |
| 9 | Sex* | Demographics & anthropometric variables | 1.35% |
| 10 | Troponin T* | Laboratory values or biomarkers | 1.33% |
| 11 | QRS duration* | Electrophysiologic variables | 1.15% |
| 12 | Cornell voltage * | Other cardiac indices | 0.96% |
| 13 | Use of anti-hypertensive medications* | Medications | 0.79% |
| 14 | Prior stroke* | Clinical factors | 0.78% |
| 15 | Race* | Demographics & anthropometric variables | 0.78% |
| 16 | Visit | Demographics & anthropometric variables | 0.61% |
| 17 | Prior heart failure | Clinical factors | 0.55% |
| 18 | Troponin I | Laboratory values or biomarkers | 0.44% |
| 19 | Blood glucose | Laboratory values or biomarkers | 0.33% |
| 20 | Systolic blood pressure | Clinical factors | 0.28% |
| 21 | Q-Q.S. pattern II, III, aVF | Electrophysiologic variables | 0.24% |
| 22 | Smoking status | Lifestyle factors | 0.21% |
| 23 | Serum creatinine | Laboratory values or biomarkers | 0.19% |
| 24 | Heart rate | Electrophysiologic variables | 0.17% |
| 25 | Education | Demographics & anthropometric variables | 0.16% |
| 26 | High-density lipoprotein cholesterol | Laboratory values or biomarkers | 0.13% |
| 27 | Age | Demographics & anthropometric variables | 0.12% |
| 28 | Hematocrit | Laboratory values or biomarkers | 0.12% |
| 29 | Urine albumin | Laboratory values or biomarkers | 0.08% |
| 30 | Body mass index | Demographics & anthropometric variables | 0.08% |
| 31 | Estimated glomerular filtration rate | Laboratory values or biomarkers | 0.08% |
| 32 | Waist circumferences | Demographics & anthropometric variables | 0.08% |
| 33 | Number of hospitalizations | Clinical factors | 0.08% |
| 34 | Diastolic blood pressure | Clinical factors | 0.06% |
| 35 | Triglycerides | Laboratory values or biomarkers | 0.04% |
| 36 | ST junction & segment depression V1-V5 | Electrophysiologic variables | 0.04% |
| 37 | Jackson field center | Demographics & anthropometric variables | 0.03% |
| 38 | Dietary quality score | Lifestyle factors | 0.03% |
| 39 | Low-density lipoprotein cholesterol | Laboratory values or biomarkers | 0.02% |
| 40 | Serum albumin | Laboratory values or biomarkers | 0.02% |
| 41 | C-reactive protein | Laboratory values or biomarkers | 0.02% |
| 42 | Atrial fibrillation | Clinical factors | 0.02% |
| 43 | Q-Q.S. pattern V1-V5 | Electrophysiologic variables | 0.02% |
| 44 | Income | Demographics & anthropometric variables | 0.02% |
| 45 | White blood cells count | Laboratory values or biomarkers | 0.02% |
| 46 | Father had coronary heart disease | Demographics & anthropometric variables | 0.01% |
| 47 | Total cholesterol | Laboratory values or biomarkers | 0.01% |
| 48 | Physical activity | Lifestyle factors | 0.01% |
| 49 | Hemoglobin | Laboratory values or biomarkers | 0.01% |
| 50 | Washington field center | Demographics & anthropometric variables | 0.01% |
| 51 | Mother had coronary heart disease | Demographics & anthropometric variables | 0.01% |
| 52 | High Amplitude R Wave Codes | Electrophysiologic variables | 0.01% |
| 53 | T Wave V1-V5 | Electrophysiologic variables | 0.01% |
| 54 | QRS axis deviation codes | Electrophysiologic variables | 0.01% |
| 55 | Minneapolis field center | Demographics & anthropometric variables | 0.005% |
| 56 | QRS transition zone | Electrophysiologic variables | 0.004% |
| 57 | Drinking status | Lifestyle factors | 0.004% |
| 58 | T Wave II, III, aVF | Electrophysiologic variables | 0.004% |
| 59 | Serum nitrogen | Laboratory values or biomarkers | 0.003% |

\*Indicate variables that together explained > 80% of the RF-SLAM prediction.
RF-SLAM: random forest statistical method for survival, longitudinal, and multivariable outcomes.

## Results

### Baseline Characteristics

Our primary study population included 15,661 participants (mean age 54.2 years; 55% female) in ARIC. Over a median follow-up of 23.5 years, there were a total of 583 adjudicated SCD cases, with 97, 93, 116, and 277 cases having occurred during the intervals of visit 1 to 2, visit 2 to 3, visit 3 to 4, and visit 4 to end of follow-up respectively. At baseline, participants who subsequently developed SCD were frequently male, self-identified as Black, had less than a high school degree, were current smokers and non-drinkers, and less physically active compared with individuals who did not develop SCD. They also had higher mean BMI and blood pressure, less favorable lipid profile, and lower kidney function when comparing with those who did not develop SCD. CHD, heart failure, stroke, and diabetes mellitus, were more prevalent among participants who developed SCD, which was also reflected by the higher percentage of participants taking medications for managing those conditions. ECG parameters among participants with SCD included higher prevalence of diagnostic Q wave, atrial fibrillation and LVH, and higher Cornell voltage (**Table 1**). The prevalence of clinical conditions at visit 4 among participants who attended visit 4 is presented at **Supplemental Table 3**. Several variables were not measured in all four visits and therefore had a high prevalence of missingness (**Supplemental Table 4**).

### RF-SLAM prediction

Using time-varying covariates from visits 1-4 of the ARIC study, the AUCs of RF-SLAM for SCD prediction at 3, 6, 9, and 25 years of follow-up were 0.83, 0.89, 0.83, and 0.77 respectively. In comparison, the AUCs for RSF were 0.82, 0.86, 0.80, and 0.75 at 3, 6, 9, and 25 years, respectively, when limiting to baseline covariates (**Figure 1A**). The predicted SCD risk by RF-SLAM with time-varying covariates ranged from 5.6×10^−2^ to 11.9 per 1000 person-years and was significantly different between SCD cases and controls, with the median of predicted risk being 3.4 per 1000 person-year ((25^th^ percentile - 75^th^ percentile, 1.7 - 5.4 per 1000 person-year) for cases and 1.1 per 1000 person-year (25^th^ percentile - 75^th^ percentile, 0.7 - 1.8 per 1000 person-year) for controls (*p*=1.6 × 10^−84^) (**Figure 1B**).

**Figure 1.**
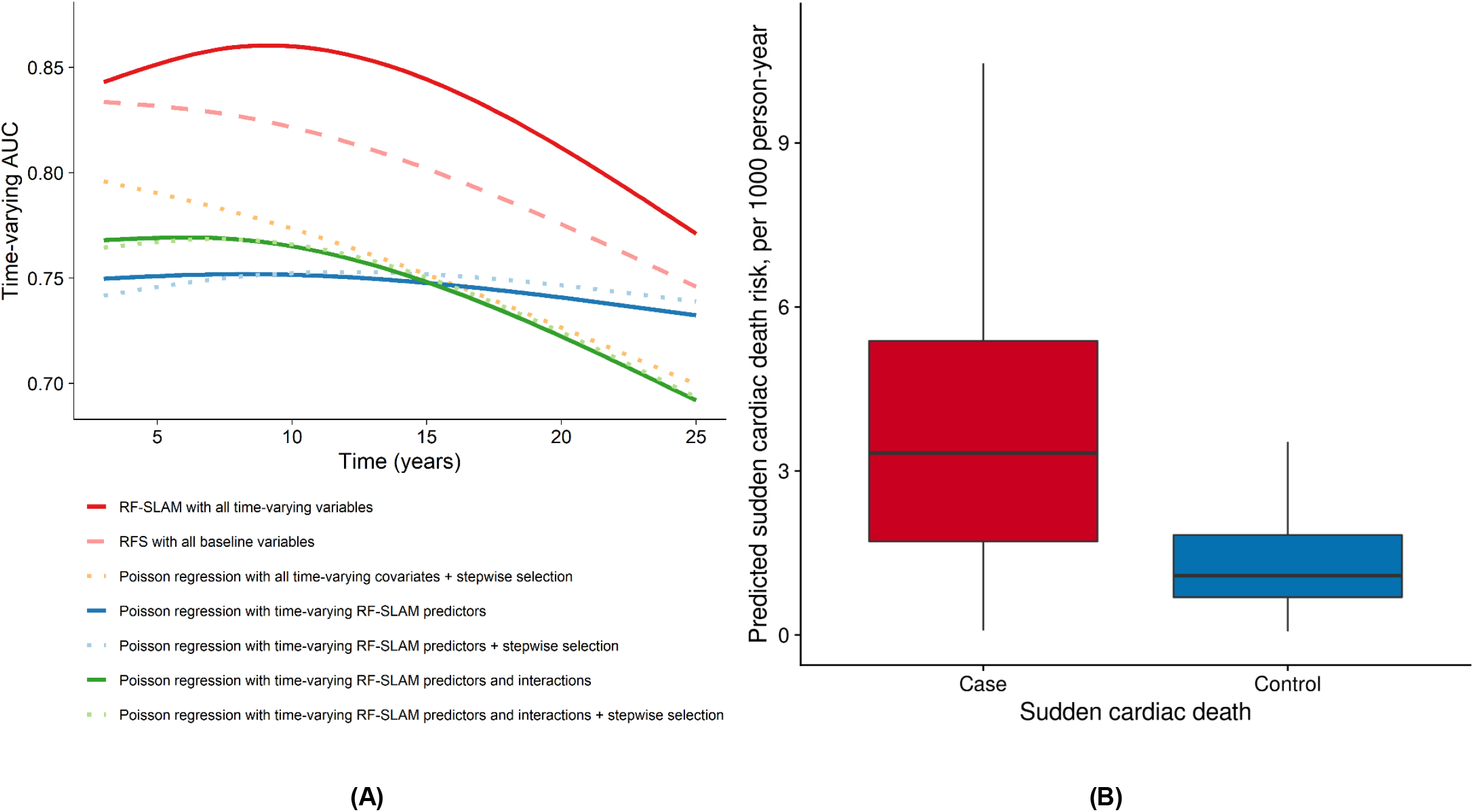
Performance of the RF-SLAM model. (A) AUC performances for predicting sudden cardiac death by RF-SLAM model incorporating time-varying covariates comparing with RFS model incorporating baseline covariates and five Poisson regression models incorporating time-varying covariates. Random forest models are in red; the model using time-varying covariates is shown as a solid line and the one using baseline covariates is shown as dashed lines with lighter color. For the five Poisson regression models, models with all candidate covariates, with predictors that accounted for > 80% of the RF-SLAM prediction, and with predictors that accounted for > 80% of the RF-SLAM prediction and their interactions are in yellow, blue, and green respectively. Models with stepwise selection are shown as solid lines and those without are shown as dashed lines with lighter colors. (B) Predicted sudden cardiac death risk per 1,000 person-year by RF-SLAM among cases and controls. Predicted risk was calculated as the mean of predicted annual risks (unit: 1,000 person-year) of all follow-up time units. Red box indicates cases and blue box indicates controls. AUC: area under the curve; RF-SLAM: random forest statistical method for survival, longitudinal, and multivariable outcomes. RFS: random forest survival.

Approximately 80% of input candidate covariates were included in the summary tree that explain 90.0 % of the RF-SLAM predictions. Top predictors that explained most of the variation included (1) clinical characteristics: CHD (variation explained: 47.9%), diabetes mellitus (12.3%), hypertension (4.9%), and number of cardiac-related hospitalization (2.2%) (2) ECG variables: Minnesota Code for T wave abnormality in any of leads I, aVL, or V6 (3.2%) and ST junction & segment depression in any of leads I, aVL, or V6 (1.2%), (3) use of medication: anti-diabetic medications (2.0%), and (4) biomarkers: NT-proBNP (3.3%), as well as sex (1.3%) (**Table 2**). Figure 2 shows the summary tree that explains 80.7% of the variation of the variation of the RF-SLAM predictions in **Figure 2** for visualizing the dependencies.

**Figure 2.**
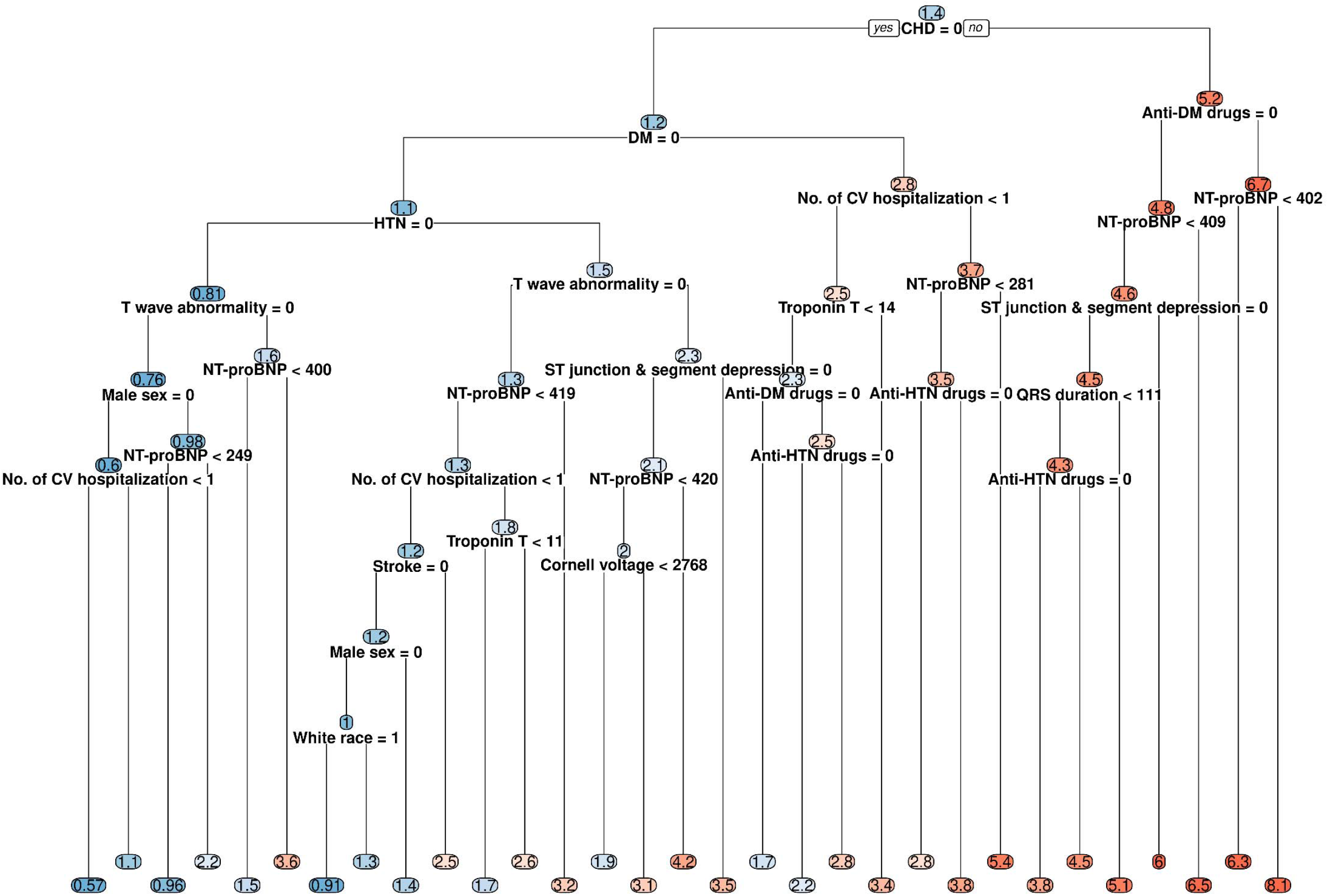
Summary tree of RF-SLAM depicting the time-varying predictors for sudden cardiac death that accounted for > 80% of the prediction. The number in each node indicate the mean predicted sudden cardiac death risk (unit: 1,000 person-year) of that node. Red nodes indicate high risk and blue nodes indicate low risk. The darker the color, the higher/lower the risk. RF-SLAM: random forest statistical method for survival, longitudinal, and multivariable outcomes. CHD: coronary heart disease, CV: cardiovascular, DM: diabetes mellitus, HTN: hypertension, NT-proBNP: N-terminal pro-brain natriuretic peptide.

### Poisson models comparison

Among all time-varying covariates, the following predictors of SCD were selected in all 5 cross-validations by stepwise regression, more specifically, a forward-backward search starting from the full model with the Akaike information criterion (AIC) for evaluating model fit: CHD, heart failure, stroke, number of all-cause hospitalization, number of cardiac-related hospitalization, three variables for ECG patterns classified by Minnesota Code (T wave in II, III, aVF, A-V conduction defect codes, and QRS transition zone), heart rate, glucose level, systolic blood pressure, diastolic blood pressure, estimated glomerular filtration rate, urinary albumin, troponin T, troponin I, fibrinogen, visit, and smoking status. Five-fold cross validation yielded AUC of 0.78, 0.83, 0.74, and 0.70 at 3, 6, 9, and 25 years of follow-up, respectively. Instead of using the stepwise variable selection, we also use the predictors selected by RF-SLAM model with and without an additional stepwise selection in the Poisson regression mode, which yielded AUC of around 0.74, 0.78, 0.73, and 0.73 at 3, 6, 9, and 25 years of follow-up, respectively, for the models with or without stepwise selection. Finally, manually adding in the top interactions identified by RF-SLAM improved the AUC at 3 and 6 years of follow-up but not for AUC at 9 and 25 years (**Figure 1A**).

For sensitivity analysis, building summary trees with bootstrapping predicted values of the RF-SLAM model yield consistent selections of predictors and percentages of total variance explained by those predictors (**Supplemental Table 5**); limiting to variables with less than 50% missingness did not change the performance of RF-SLAM prediction (AUC at 3 years: 0.83, 6 years: 0.88, 9 years: 0.83, 25 years: 0.77) nor the selection of predictors (**Supplemental Figure 1 and Supplemental Table 6**); excluding participants with prevalent CHD at baseline resulted in slight drop in the performance (AUC at 3 years: 0.81, 6 years: 0.87, 9 years: 0.78, 25 years: 0.75) and the predictors other than CHD remained similar (**Supplemental Figure 1 and Supplemental Table 7**).

## Discussion

In this community-based cohort of 15,663 middle-aged adults, we derived a prediction model for SCD with a novel ML approach, RF-SLAM using a large number of time-varying covariates. Participants who developed SCD had statistically significant higher predicted risk of SCD than those who did not. This model substantially outperformed a random forest survival model with only baseline covariates as well as Poisson regression models with time-varying predictors selected through stepwise regression. Prior CHD was identified as the top predictor for SCD, explaining 47.9% of the total phenotypic variance. The combination of these time-varying data and ML methods that accommodate dynamic data can contribute to improved risk stratification for SCD in the generally healthy population and thus aid in targeted primary prevention strategies for high-risk individuals.

Our study highlighted the importance of the dynamic dependency of time-varying risk factors on SCD prediction. Leveraging novel ML approach that incorporates large number of time-varying predictor variables, we generated models that providing continuous a gradient of predicted SCD risk with significant differentiation between those who ultimately have versus do not have an SCD. This model demonstrated better performance than both using same ML approach but only inputting baseline variables and using traditional statistical regression analysis with stepwise selection of time-varying variables.

A single summary decision tree facilitates the portability and interpretability of complex clinical ML algorithms. The “black box” algorithms of ML methods can limit their clinical or epidemiologic utility. To surmount this limitation, we approximated the predictions from the complex algorithm into a single summary decision tree with represents the sequential subsetting that produces groups of differential risk and mimics clinical reasoning. In this way, we (1) identified the key predictors as well as calculated the variation in the risks among people explained by each predictor, which can be easily used as predictors by other comparable general population cohorts; and (2) visualize the interactions among predictors as reflected in the sequences of splitting variables, which facilitate understanding the dependency among risk factors. Top time-varying predictors identified using RF-SLAM including CHD, diabetes mellitus, hypertension, NT-proBNP, and T wave abnormality in any of leads I, aVL, and V6,, which are consistent with existing epidemiological findings.^9,33,34^ Among them, prior CHD alone explained 47.9% of total variation of the RF-SLAM prediction, which corroborate with the fact that approximately 50% of SCD victims have prior diagnosis of heart disease.^1^

Our identification of key predictors of SCD in a community-based cohort can better inform the prevention and management strategy for the general population. In our study, in addition to CHD, diabetes, hypertension along with their corresponding biomarkers and medications, explained a large proportion of the total variation of SCD prediction. Diabetes and hypertension are widely recognized modifiable cardiovascular risk factors. Furthermore, our study indicates that clinical surveillance may be appropriate for patients with CHD, diabetes, and hypertension for risk of SCD. For instance, monitoring could include serial electrocardiograms to assess features such as T wave abnormalities and ST junction & segment depression, as well as cardiac biomarkers, such as NT-proBNP, which were selected as top predictors for SCD in our study. A multitude of high-risk features may also prompt further risk stratification with echocardiography among asymptomatic patients.

Other features of the ML approach favor it use over classical regression methods. In random forest, each tree is built from a bootstrap sample of the observations in the training data. As a result of the bootstrapping, roughly one in three observations is randomly left out in a particular bootstrap. We can predict these “out-of-bag” observations from the model fit to the “in-bag” values and obtain a cross-validated estimate of prediction error along the way. Another key feature is that missing data imputation can be integrated within random forests but must be handled as separate steps for regression analysis.^25^ Particularly in our study, through randomly drawing from the non-missing “in-bag” data within the current node during the tree growing process, RF-SLAM demonstrated its robustness to missingness, a common and important issue in real-world longitudinal data, compared to conventional methods.

Several limitations should be considered when interpreting our findings. First, ECG has long offered valuable insights into cardiac health and disease.^35,36^ Several ECG abnormalities have been associated with SCD risk^37^ and our models also selected some of them as predictors. We used the Minnesota code for ECG variables while recent advances in deep-learning convolutional neural networks (CNNs) have been used to extract information beyond that being captured by Minnesota code.^38^ We will incorporate these additional ECG features generated through CNNs when they become available in the study population. Second, there is a time gap between visit 4 and the end of follow-up of our study, limiting the incorporation of features during this interval and likely contributing to the modest AUC decline during this interval. Third, for important clinical events predictors such as CHD and heart failure, we updated them just at each ARIC visit but not for events during each interval. Fourth, some candidate covariates in our study were not measured in all four visits resulting in substantial missingness. These include important cardiac biomarkers NT-proBNP and troponin T that have been associated with risk of multiple cardiovascular events including SCD.^39-44^ To address this, we conducted sensitivity analysis with excluding those covariates, and observed results consistent with our main analysis. Finally, cross-validation was performed in the diverse ARIC cohort and future research is needed to confirm generalizability in external cohorts.

In conclusion, our study highlighted the improved prediction for SCD of using a novel machine-learning approach with time-varying predictors, as well as the feasibility of applying this approach in large cohorts and biobanks. Our findings allow identification of higher-risk individuals appropriate for targeted interventions designed to reduce the burden of SCD in the general population.

## Data Availability

All data produced in the present study are available upon reasonable request to the authors.

## Acknowledgements

The authors thank the staff and participants of the ARIC study for their important contributions.

