## Supplemental material for "Machine Learning for Sudden Cardiac Death Prediction in the Atherosclerosis Risk in Communities Study"

**Supplemental Table 1. Definition and measurement details of laboratory values or biomarkers.**

|  | **Definitions** | **Measurements** | | | |
| --- | --- | --- | --- | --- | --- |
|  |  | **Visit 1** | **Visit 2** | **Visit 3** | **Visit 4** |
| **C-reactive protein (CRP)** | Plasma levels of high sensitivity CRP were determined by an immunophelometric assay (Siemens Healthcare Diagnostics, Deerfield, IL). |  | ✓ |  | ✓ |
| **White blood cells (WBC) count** | WBC count was determined by automated particle Coulter Counters based on WBCs retrieved from whole anti-coagulated blood within 24 hours after venipuncture in local hospital hematology laboratories. | ✓ | ✓ | ✓ | ✓ |
| **Hematocrit** | Hematocrit levels were calculated from the measurement of red blood cells and either the calculated erythrocyte mean cell volume using Coulter counter (Coulter Diagnostics, Hialeah, FL) or pattern of light scattering using Hemalog H-6000 (Technicon Corporation, Tarrytown, NY). | ✓ | ✓ | ✓ | ✓ |
| **Hemoglobin** | Hemoglobin was measured using automated hematology analyzers, Coulter S+IV (Beckman Coulter, Fullerton, CA) and Technicon H‐6000 (Technicon Corporation, Tarrytown, NY). | ✓ | ✓ |  | ✓ |
| **N-terminal pro-brain natriuretic peptide (NT-proBNP)** | Plasma NT-proBNP levels were measured in stored plasma samples using an electrochemiluminescent immunoassay on an automated Cobas e411 analyzer (Roche Diagnostics, Indianapolis, IN). The lower limit of detection for this assay is 5 pg/ml. |  | ✓ | ✓ | ✓ |
| **Troponin I** | High sensitivity troponin I (hs-TnI) levels were measured using a chemiluminescent immunoassay, Architect Stat Troponin-I (Abbott Laboratories, Abbott Park, IL) on an automated chemistry analyzer, Architect i 2000sr (Abbott Laboratories, Abbott Park, IL). The limit of detection of the hs-TnI assay was 1.2 ng/L. |  |  |  | ✓ |
| **Troponin T** | High sensitivity troponin T (hs-TnT) levels were measured using a high-sensitivity assay implemented on an automated Cobas e411 analyzer, Elecsys Troponin T (Roche Diagnostics, Indianapolis, IN). The limit of detection of the hs-TnT assay was 5 ng/L and undetectable levels were assigned a value of 5 ng/L. |  | ✓ | ✓ | ✓ |
| **Fibrinogen** | Plasma fibrinogen levels were determined using the thrombin time titration method with reagents (General Diagnostics Organon Technica Co., Morris Plains, NJ). | ✓ |  |  |  |
| **Total cholesterol** | Total cholesterol levels were measured using enzymatic methods with reagents (Boehringer-Mannheim Biochemical, Indianapolis, IN) and adapted for analysis using the Cobas-Bioanalyzer (Roche, Montclair, NJ). | ✓ | ✓ | ✓ | ✓ |
| **High-density lipoprotein (HDL) cholesterol** | HDL cholesterol levels were determined enzymatically after dextran sulfate-Mg^2+^ precipitation of other lipoproteins. | ✓ | ✓ | ✓ | ✓ |
| **Low-density lipoprotein (LDL) cholesterol** | LDL cholesterol levels were estimated with the Friedewald formula. | ✓ | ✓ | ✓ | ✓ |
| **Triglycerides** | Triglycerides levels were measured using enzymatic methods with reagents (Boehringer-Mannheim Biochemical, Indianapolis, IN) and adapted for analysis using the Cobas-Bioanalyzer (Roche, Montclair, NJ). | ✓ | ✓ | ✓ | ✓ |
| **Serum creatinine** | Serum creatinine levels were measured by the modified kinetic Jaffé method, standardized to the National Institute of Standards and Technology standard, and calibrated to an isotope dilution mass spectrometry-traceable reference method. | ✓ | ✓ | ✓ |  |
| **Estimated glomerular filtration rate (eGFR)** | eGFR was calculated using the 2009 Chronic Kidney Disease Epidemiology Collaboration (CKD-EPI) creatinine equation | ✓ | ✓ | ✓ |  |
| **Serum albumin** | Serum albumin concentrations were measured with a Coulter DACOS instrument (Coulter Diagnostics, Hialeah, FL) using Coulter's bromcresol green colorimetric assay. | ✓ |  | ✓ | ✓ |
| **Urine albumin** | Concentrations of urinary albumin were determined separately by nephelometry and the Jaffé method, respectively using spot urine samples. |  |  |  | ✓ |
| **Blood glucose** | Serum glucose was assayed by a hexokinase/glucose-6-phosphate dehydrogenase method. | ✓ | ✓ | ✓ | ✓ |
| **Serum sodium (Na)** | Serum Na levels were assessed with a Coulter DACOS analyzer (Coulter Instruments, Hialeah, FL) using a direct ion-selective electrode. | ✓ | ✓ |  |  |
| **Serum potassium (K)** | Serum K levels were measured on a Coulter DACOS analyzer (Coulter Instruments, Hialeah, FL) using a direct ion-selective electrode. | ✓ | ✓ |  |  |
| **Blood urea nitrogen (BUN)** | Serum BUN concentrations were determined by an Architect Ci8200 automatic analyzer (Abbott Laboratories. Abbott Park, IL) with Abbott diagnostic reagents following standard experiment procedures provided by the manufacturer. | ✓ |  |  |  |
| **Serum magnesium (Mg)** | Serum Mg levels were measured based on the procedure of Gindler and Heth using the metallochromic dye, Calmagite (1-(1-hydroxy-4-methyl-2-phenylazo)-2-napthol-4sulfonic acid). | ✓ | ✓ |  |  |

**Supplemental Table 2. Comparisons between models used for prediction.**

|  | **RF-SLAM** | **Random forest survival** | **Poisson regression model with all candidate variables** | **Poisson regression model with RF-SLAM predictors** | **Poisson regression model with RF-SLAM predictors and interactions** |
| --- | --- | --- | --- | --- | --- |
| **Data** | Time-varying covariates from four visits | Baseline covariates | Time-varying covariates from four visits | | |
| **Underlying model** | Poisson regression log-likelihood for splitting rates | Log-rank splitting | Poisson regression with offset | | |
| **Predictors selection** | Splitting variables maximize the difference in children nodes | | Stepwise regression (AIC) | | |
| **Missing data** | Imputation by randomly drawing from non-missing data | | Multivariate Imputation by Chained Equations (MICE) | | |
| **Validation** | Out-of-bag predictions | | 5-fold cross validation | | |
| **Covariates** | All covariates | | | Predictors selected in RF-SLAM model | |
| **Interaction/non-linearity** | Captured in regression tree | | Not captured; assuming additive effects | | Manually add interactions identified in RF-SLAM model |

ARIC: Atherosclerosis Risk in Communities Study; RF-SLAM: random forest statistical method for survival, longitudinal, and multivariable outcomes.

**Supplemental Table 3.  Characteristics of clinical conditions at visit 4 of Atherosclerosis Risk in Communities (ARIC) study participants.**

| **Metric, N (%)** | **No SCD (N = 11,277)** | **SCD (N = 277)** | **P-value** |
| --- | --- | --- | --- |
| Coronary heart disease | 854 (7.7) | 89 (33.1) | <0.001 |
| Heart failure | 574 (5.1) | 48 (17.3) | <0.001 |
| Stroke | 245 (2.2) | 21 (7.6) | <0.001 |
| Diabetes mellitus | 1266 (11.2) | 82 (29.6) | <0.001 |
| Hypertension | 5304 (47.3) | 184 (66.9) | <0.001 |
| Atrial fibrillation | 105 (0.9) | 4 (1.5) | 0.574 |

**Supplemental Table 4. Candidate covariates with missingness > 10% of total observations.**

|  | **% of missingness of total observations** |
| --- | --- |
| **Troponin I** | 80% |
| **Urine albumin** | 79% |
| **Troponin T** | 74% |
| **Fibrinogen** | 72% |
| **Serum nitrogen** | 71% |
| **N-terminal pro-brain natriuretic peptide** | 56% |
| **C-reactive protein** | 55% |
| **Serum albumin** | 53% |
| **Dietary quality score** | 49% |
| **Physical activity (intensity)** | 48% |
| **Physical activity (meeting AHA recommendations or not)** | 48% |
| **Presence of diagnostic Q-wave** | 45% |
| **Serum magnesium** | 45% |
| **Serum sodium** | 45% |
| **Serum potassium** | 45% |
| **white blood cells count** | 28% |
| **Hematocrit** | 28% |
| **Serum creatinine** | 24% |
| **Estimated glomerular filtration rate** | 24% |
| **Carotid intima-media thickness** | 22% |
| **Use of anti-diabetic medications** | 11% |

AHA: American Heart Association

**Supplemental Table 5. Top 10 predictors that explained the largest variation of RF-SLAM prediction for sudden cardiac death selected through regressing the bootstrapped predicted risk on candidate variables.**

| **Bootstrap 1** | **Bootstrap 2** | **Bootstrap 3** | **Bootstrap 4** | **Bootstrap 5** | **Bootstrap 6** | **Bootstrap 7** | **Bootstrap 8** | **Bootstrap 9** | **Bootstrap 10** |
| --- | --- | --- | --- | --- | --- | --- | --- | --- | --- |
| Prior coronary heart disease (48%) | Prior coronary heart disease (48.5%) | Prior coronary heart disease (45.3%) | Prior coronary heart disease (49.9%) | Prior coronary heart disease (51.5%) | Prior coronary heart disease (47.4%) | Prior coronary heart disease (46.9%) | Prior coronary heart disease (48.4%) | Prior coronary heart disease (49.7%) | Prior coronary heart disease (48.7%) |
| Diabetes mellitus (13.8%) | Diabetes mellitus (12.5%) | Diabetes mellitus (13.7%) | Diabetes mellitus (12.2%) | Diabetes mellitus (14.2%) | Use of anti-diabetic medications (14.2%) | Diabetes mellitus (11.4%) | Use of anti-diabetic medications (14.4%) | Use of anti-diabetic medications (12.5%) | Diabetes mellitus (12.7%) |
| Hypertension (5%) | Hypertension (5%) | Hypertension (4.9%) | Hypertension (5%) | Hypertension (5%) | Hypertension (5.4%) | Hypertension (5.2%) | Hypertension (5.2%) | Hypertension (5%) | Hypertension (5%) |
| N-terminal pro-brain natriuretic peptide (3.5%) | N-terminal pro-brain natriuretic peptide (3.8%) | T wave abnormality in any of leads I, aVL, and V6 (3.8%) | T wave abnormality in any of leads I, aVL, and V6 (4.3%) | T wave abnormality in any of leads I, aVL, and V6 (3.3%) | T wave abnormality in any of leads I, aVL, and V6 (3.6%) | T wave abnormality in any of leads I, aVL, and V6 (3.7%) | N-terminal pro-brain natriuretic peptide (3.8%) | T wave abnormality in any of leads I, aVL, and V6 (3.5%) | T wave abnormality in any of leads I, aVL, and V6 (3.4%) |
| T wave abnormality in any of leads I, aVL, and V6 (3.2%) | T wave abnormality in any of leads I, aVL, and V6 (2.7%) | Number of cardiac-related hospitalization (3.2%) | N-terminal pro-brain natriuretic peptide (2.2%) | N-terminal pro-brain natriuretic peptide (2.7%) | N-terminal pro-brain natriuretic peptide (2.7%) | N-terminal pro-brain natriuretic peptide (3.3%) | T wave abnormality in any of leads I, aVL, and V6 (3.3%) | N-terminal pro-brain natriuretic peptide (3.1%) | Sex (1.3%) |
| Number of cardiac-related hospitalization (2.2%) | Number of cardiac-related hospitalization (2%) | N-terminal pro-brain natriuretic peptide (2%) | Number of cardiac-related hospitalization (2%) | ST junction & segment depression in any of leads I, aVL, or V6 (2.2%) | Number of cardiac-related hospitalization (2%) | Troponin T (1.9%) | Number of cardiac-related hospitalization (2.4%) | Diabetes mellitus (2.2%) | Number of cardiac-related hospitalization (2.5%) |
| Sex (1.3%) | ST junction & segment depression in any of leads I, aVL, or V6 (1.2%) | Sex (1%) | Troponin T (1.6%) | Number of cardiac-related hospitalization (1.1%) | Sex (1.2%) | Use of anti-diabetic medications (1.8%) | ST junction & segment depression in any of leads I, aVL, or V6 (1.3%) | Number of cardiac-related hospitalization (1.9%) | QRS duration (0.6%) |
| ST junction & segment depression in any of leads I, aVL, or V6 (1.3%) | Sex (1.1%) | Troponin T (0.8%) | Sex (1.1%) | Sex (1%) | Cornell voltage (1.1%) | Number of cardiac-related hospitalization (1.5%) | Sex (1.2%) | ST junction & segment depression in any of leads I, aVL, or V6 (1.4%) | ST junction & segment depression in any of leads I, aVL, or V6 (1.7%) |
| Troponin T (0.8%) | Prior stroke (0.4%) | ST junction & segment depression in any of leads I, aVL, or V6 (0.7%) | Prior stroke (0.9%) | Troponin T (0.8%) | ST junction & segment depression in any of leads I, aVL, or V6 (1.1%) | Sex (0.9%) | Troponin T (0.6%) | Sex (1%) | Prior stroke (0.4%) |
| Prior stroke (0.6%) | Cornell voltage (0.3%) | Cornell voltage (0.6%) | Race (0.3%) | QRS duration (0.6%) | Troponin T (0.9%) | Cornell voltage (0.9%) | Use of anti-hypertensive medications (0.4%) | Cornell voltage (0.9%) | Cornell voltage (0.4%) |

RF-SLAM: random forest statistical method for survival, longitudinal, and multivariable outcomes.

**Supplemental Table 6. Top 10 predictors that explained the largest variation of RF-SLAM prediction for sudden cardiac death with excluding candidate variables with > 50% missingness.**

|  | **Variable** | **Category** | **% of total variation explained** |
| --- | --- | --- | --- |
| 1 | Prior coronary heart disease | Clinical factors | 42.83% |
| 2 | Diabetes mellitus | Clinical factors | 13.65% |
| 3 | Hypertension | Clinical factors | 5.60% |
| 4 | T wave abnormality in any of leads I, aVL, and V6 | Electrophysiologic variables | 4.63% |
| 5 | ST junction & segment depression in any of leads I, aVL, or V6 | Electrophysiologic variables | 2.70% |
| 6 | Number of cardiac-related hospitalization | Clinical factors | 2.45% |
| 7 | QRS duration | Electrophysiologic variables | 2.37% |
| 8 | Sex | Demographics & anthropometric variables | 1.22% |
| 9 | Use of anti-hypertensive medications | Medications | 1.20% |
| 10 | Cornell voltage | Other cardiac indices | 0.72% |

RF-SLAM: random forest statistical method for survival, longitudinal, and multivariable outcomes.

**Supplemental Table 7. Top 10 predictors that explained the largest variation of RF-SLAM prediction for sudden cardiac death among participants without baseline coronary heart disease.**

|  | **Variable** | **Category** | **% of total variation explained** |
| --- | --- | --- | --- |
| 1 | Diabetes mellitus | Clinical factors | 23.47% |
| 2 | Hypertension | Clinical factors | 11.05% |
| 3 | N-terminal pro-brain natriuretic peptide | Laboratory values or biomarkers | 6.90% |
| 4 | T wave abnormality in any of leads I, aVL, and V6 | Electrophysiologic variables | 5.38% |
| 5 | Troponin I | Laboratory values or biomarkers | 3.31% |
| 6 | Number of cardiac-related hospitalization | Clinical factors | 3.11% |
| 7 | Cornell voltage | Other cardiac indices | 2.52% |
| 8 | ST junction & segment depression in any of leads I, aVL, or V6 | Electrophysiologic variables | 1.97% |
| 9 | Race | Demographics & anthropometric variables | 1.95% |
| 10 | Sex | Demographics & anthropometric variables | 1.91% |

RF-SLAM: random forest statistical method for survival, longitudinal, and multivariable outcomes.

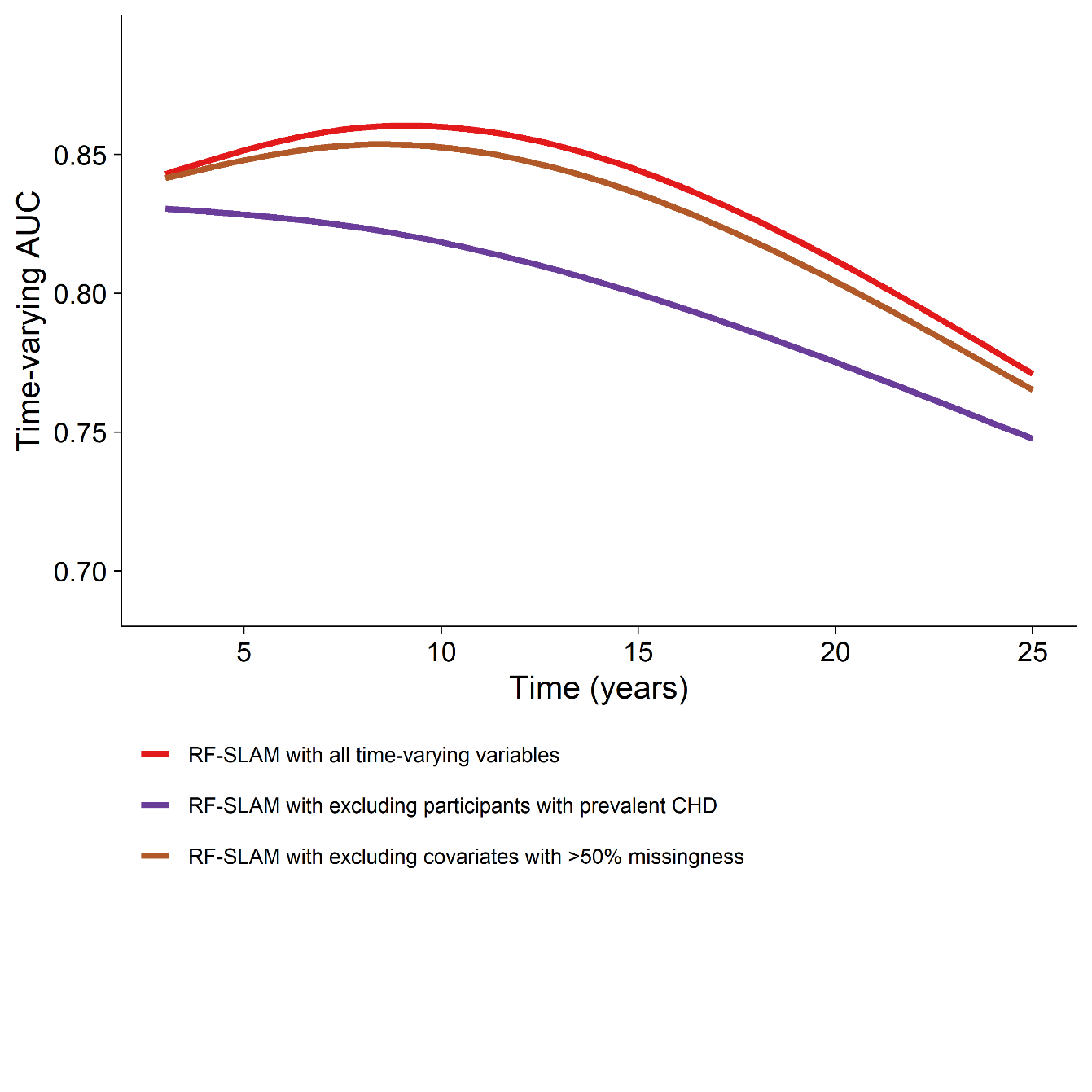

**Supplemental Figure 1.** AUC performances for predicting sudden cardiac death by RF-SLAM model including all participants and all time-varying covariates (main model) comparing with excluding participants with coronary heart disease at baseline or excluding covariates with >50% missingness. Main model is in red; the model with excluding participants with prevalent coronary heart disease is in brown; the model with excluding participants with covariates with >50% missingness is in purple. AUC: area under the curve; RF-SLAM: random forest statistical method for survival, longitudinal, and multivariable outcomes.
